# Global research hotspots and frontier trends of epigenetic modifications in autoimmune diseases: a bibliometric analysis from 2012 to 2022

**DOI:** 10.1101/2023.07.05.23292268

**Authors:** Xiang Gao, Xin Huang, Yehui Wang, Sheng Sun, Tao Chen, Yongxiang Gao, Xiaodan Zhang

**Affiliations:** Hospital of Chengdu University of Traditional Chinese Medicine, Chengdu 610072, Sichuan Province, China; International Education College, Chengdu University of Traditional Chinese Medicine, Chengdu 610075, Sichuan Province, China; Chengdu University of Traditional Chinese Medicine, Chengdu, 610075, Sichuan Province, China

**Author notes:** Correspondence: Xiaodan Zhang: Yongxiang Gao. These authors contributed equally to this work (Xiang Gao, Xin Huang).

**Keywords:** Autoimmune diseases, Epigenetic Modification, Bibliometrics;DNA methylation;VOSviewer

## Abstract

**Background:** Epigenetics plays a significant role in the pathogenesis of autoimmune diseases (ADs) and has garnered considerable attention in related research fields. Recent studies have shown substantial progress in understanding the association between epigenetics and autoimmune diseases. However, there is a lack of comprehensive bibliometric analysis in this research area.

**Objective:** This article aims to present the current status and hot topics of epigenetic research in ADs from a bibliometric perspective, as well as explore the frontier hotspots and trends in epigenetic studies related to ADs.

**Methods:** This study collected 1,870 epigenetic records related to autoimmune diseases from the Web of Science Core Collection (WoSCC) database, spanning from 2012 to 2021. Analysis of regions, institutions, journals, authors, and keywords was conducted using CiteSpace, VOSviewer, and the R package "bibliometrix" to predict the latest trends in epigenetic research relevant to autoimmune diseases.

**Results:** The number of epigenetic publications related to autoimmune diseases has been increasing annually. The United States has played a major role in this field, contributing over 45.9% of publications and leading in terms of publication volume and citation counts. Central South University emerged as the most active institution, contributing the highest number of publications. Frontiers in Immunology is the most popular journal in this field, publishing the most articles, while the Journal of Autoimmunity is the most co-cited journal. Lu QJ is the most prolific author, and Zhao M is the most frequently co-cited author. "Immunology" serves as a broad representative of epigenetic research in ADs. Hot topics in the field of epigenetic modifications associated with autoimmune diseases include "regulatory T cells (Treg)," "rheumatoid arthritis," "epigenetic regulation," "cAMP-responsive element modulator alpha," "cell-specific enhancer," "genetic susceptibility," and "systemic lupus erythematosus." Furthermore, the study discusses the frontiers and existing issues of epigenetic modifications in the development of autoimmune diseases.

**Conclusion:** This study provides a comprehensive overview of the knowledge structure and developmental trends in epigenetic research related to autoimmune diseases over the past 11 years. The research findings summarize and clarify the forefront and future trends in studying the association between epigenetics and autoimmune diseases, serving as a reference for scholars engaged in exploring the relationship between epigenetics and autoimmune diseases

**Key Points:** This article presents the current status and hot topics of epigenetic research in ADs from a bibliometric perspective, as well as explore the frontier hotspots and trends in epigenetic studies related to ADs.Some potential future directions for epigenetic research in the context of autoimmune diseases include elucidating the mechanisms of epigenetic regulation in disease occurrence and development through annotation of specific genomic regions, with a greater emphasis on targeting microRNAs through mimics or inhibitors and cell-based therapies for the treatment of autoimmune diseases. These approaches hold great value and promise in regulating various aspects of human autoimmune diseases. It can be predicted that further collaboration between authors, institutions, and countries in the future will accelerate the development of epigenetic research related to autoimmune diseases

## Introduction

Autoimmune diseases are a group of disorders caused by the abnormal attack of the immune system on the body’s own tissues and organs ^[1,2]^. Currently, there are over 80 known autoimmune diseases, including autoimmune disorders, Crohn’s disease, hyperthyroidism, multiple sclerosis, and others ^[3]^. The incidence of autoimmune diseases is increasing worldwide, with women being more susceptible than men and it being a leading cause of death in young women ^[4]^. The exact mechanisms underlying autoimmune diseases are not fully understood, but it is believed to involve genetic factors, environmental factors, and immune dysregulation ^[5,6]^.

Diagnosis and treatment of autoimmune diseases pose several challenges. Firstly, the clinical manifestations of autoimmune diseases are diverse and atypical, making laboratory tests insensitive and nonspecific, leading to delays in diagnosis or misdiagnosis. This can affect the effectiveness of early treatment and prognosis. Therefore, there is a need to identify new biomarkers to improve early diagnostic capabilities for autoimmune diseases. Secondly, a cure for autoimmune diseases remains elusive. Current treatment options can only control the activity of the immune response, alleviate tissue damage, and slow disease progression, but they cannot eliminate the root cause of the autoimmune response. Additionally, many patients develop drug resistance or are unable to tolerate the side effects of treatment. Hence, exploring new therapeutic strategies to completely eliminate the autoimmune response is necessary.

Epigenetic modifications are molecular mechanisms that regulate gene expression without altering the DNA sequence. Examples include DNA methylation, histone modifications, and non-coding RNA ^[7]^. Epigenetic modifications can reflect the cellular state, function, and the impact of environmental factors ^[8]^. Therefore, they hold diagnostic and therapeutic value in autoimmune diseases. Epigenetic modifications can serve as potential biomarkers for autoimmune diseases, aiding in early diagnosis, classification, prognosis assessment, and monitoring of drug responses. For example, in patients with autoimmune diseases, the level of DNA methylation is negatively correlated with disease activity and severity, serving as an indicator of disease status ^[9]^. Furthermore, epigenetic modifications can also be targeted for therapeutic intervention in autoimmune diseases by altering the levels of epigenetic modifications to regulate immune cell function and the activity of the autoimmune response. This therapeutic approach is known as "epigenetic therapy" and offers advantages such as reversibility, selectivity, and specificity ^[10]^.

Bibliometric analysis is a scientific method that utilizes statistical techniques to analyze publications such as books and articles, particularly in scientific content. It can help us understand the development trends in a specific field, identify emerging research areas, recognize potential research collaborators and journals, and assess the quality and impact of literature. In this article, we will conduct a visual analysis and summary of the relevant studies on epigenetics in autoimmune diseases using bibliometric analysis. This will showcase the current research hotspots and frontier trends of epigenetics in autoimmune diseases, facilitating a better understanding of the role of epigenetics in the mechanisms, diagnosis, and treatment of autoimmune diseases. Various software tools can be used for bibliometric analysis, such as Citespace, VOSviewer, and R-Bibliometric. These tools can generate visualizations such as network maps, overlay maps, and timeline graphs to display information about the countries/regions, institutions, authors, journals, references, keywords, and more in the literature data.

## Methods

### Data Source

The original data for this study was obtained from the Web of Science Core Collection, which is the largest and most authoritative database in the world. Two researchers independently conducted the search process, limiting the publication date from January 1, 2012, to December 31, 2022. The search query used was as follows: TS=("epigenetic" or "epigenomic" or "epigenetics") AND TS=("autoimmunity" or "autoimmune"). The search was performed on May 29, 2023, and a total of 1,955 articles were retrieved. The following content was excluded: book chapters (38), editorial materials (28), conference papers (21), conference abstracts (14), online publications (6), revisions (3), letters (2), book reviews (1), resulting in a final selection of 1,870 papers and reviews. The language was limited to English, excluding 26 articles. The data was exported in plain text format and named as "download." See Figure 1 for details.

### Visualization Analysis

For the visualization analysis of the data, we utilized three software tools: CiteSpace (version 6.1.6R) ^[11,12]^, VOSviewer (version 1.6.18) ^[13]^, and R-Bibliometric ^[14]^. These tools allow us to generate visual effects such as network maps, overlay maps, and timeline graphs, demonstrating authors, institutions, countries/regions, journals, articles, keywords, subject terms, and references. They also facilitate bibliometric analysis such as co-authorship, co-occurrence, and co-citation analysis ^[16–18]^. In this study, we employed Citespace ^[19]^ to perform cluster analysis, burst detection, timeline analysis, and journal co-occurrence analysis of references. The relationship between citing journals and cited journals was represented by overlay dual-map visualization ^[20]^. VOSviewer was used to create network visualizations of countries, institutions, co-authorship, co-citation authors, and keywords. VOSviewer focuses on graphical descriptions in bibliometric studies ^[21]^. Each node on the VOSviewer map corresponds to a specific parameter such as author, institution, or country. The size of the nodes represents the number of publications, citation counts, or occurrence frequencies. The color indicates the clusters to which nodes and lines belong. Connections are represented by lines connecting nodes ^[22]^. Additionally, we used R-Bibliometric software to visualize the publication output maps by country, international collaboration, and three-field maps, among others.

### Ethical Approval

Ethical approval was not required for this study because the information used was publicly available and did not involve any interaction with human subjects.

## Results

### Overall Analysis of Global Publications

A total of 1,870 articles on epigenetic modifications in autoimmune diseases were included in this study, covering the period from 2012 to 2022. Table 1 shows the number of articles, citation counts, and average citation counts per year. The average citation count is a metric used to measure the quality and impact of articles in bibliometrics. From Table 1, it can be observed that articles published in 2013 and 2014 had the highest average citation counts, with 77.98 and 73.33, respectively. The highest average citation count per year was observed in 2019, with 8.29. Figure 2(A) illustrates the trends in the number of articles and citation counts per year, indicating an overall increasing trend in both, with a peak in 2022, with 261 articles (13.96% of the total) and 18,567 citations. This suggests that epigenetic research in autoimmune diseases has gained widespread attention and citation.

**Table 1.** Global publication output and citation trend of epigenetic modifications in autoimmune diseases

| Year | Mean TC per Article | Publications | Mean TC perYear | Total Citations |
| --- | --- | --- | --- | --- |
| 2012 | 66.03 | 94 | 5.5 | 179 |
| 2013 | 77.98 | 108 | 7.09 | 886 |
| 2014 | 73.33 | 122 | 7.33 | 1,821 |
| 2015 | 66.46 | 146 | 7.38 | 2,821 |
| 2016 | 48.48 | 133 | 6.06 | 3,944 |
| 2017 | 41.61 | 174 | 5.94 | 5,138 |
| 2018 | 36.8 | 178 | 6.13 | 6,031 |
| 2019 | 41.47 | 214 | 8.29 | 7,662 |
| 2020 | 22.99 | 206 | 5.75 | 10,368 |
| 2021 | 10.74 | 234 | 3.58 | 13,204 |
| 2022 | 3.31 | 261 | 1.66 | 18567 |

Furthermore, we conducted a statistical analysis of the number of articles and citation counts from different countries/regions. Figure 2(B) displays a map of publication output by country/region, indicating that North America, Asia, and Europe are the major contributors in this field. Figures 2(C) and2(D) present the annual publication counts and total publication counts for the top ten countries/regions. The United States leads in this field, with a total of 522 publications (27.91% of the total) and 28,267 citations. China ranks second, with 337 publications (18.02% of the total) and 9,691 citations. Italy ranks third, with 148 publications (7.91% of the total) and 4,088 citations. These results reflect the global distribution and development of research on the relationship between epigenetics and autoimmune diseases.

### Country/Region Analysis

According to the statistics, a total of 76 countries participated in research related to epigenetics and autoimmune diseases from 2012 to 2022. The top 10 most cited countries are shown in Table 2, and similarly, Figures 3A and 3B show the total and average number of citations for the top 20 countries, respectively..The United States contributed the most with 28,267 citations and an average citation count of 54.2. China followed with 9,691 citations and an average of 28.8, and Italy with 4,088 citations and an average of 27.6. In terms of international collaboration between countries/regions, as shown in Figure 3C, the United States had collaborations with many countries, followed by China and the United Kingdom. However, the level of collaboration between other countries remained relatively low. Additionally, TLS (Total Link Strength) represents the strength of the connections between linked nodes, indicating the level of international collaboration. Using VOSviewer, we analyzed the global collaboration network with a minimum threshold of 5 papers. As shown in Figure 3D, the top 5 countries in terms of TLS were the United States (TLS=4,502), China (TLS=3,050), Italy (TLS=1,530), the United Kingdom (TLS=1,246), and Germany (TLS=924). SCP represents the number of publications co-authored by authors from the same country, while MCP represents the number of publications co-authored by authors from different countries. The MCP ratio can be considered as a measure of global collaboration, with higher values indicating stronger international collaboration. Combining Table2 and Figure3E, Germany had the highest international collaboration ratio (0.45), followed by the United Kingdom and France.

**Table 2.** Productive countries/regions related to epigenetic modifications in autoimmune diseases

| Country | Articles | SCP | MCP | MCP-Ratio | Citations | Average Article Citations | TLS |
| --- | --- | --- | --- | --- | --- | --- | --- |
| USA | 522 | 410 | 112 | 0.21 | 28267 | 54.2 | 4502 |
| CHINA | 337 | 241 | 96 | 0.28 | 9691 | 28.8 | 3050 |
| ITALY | 148 | 101 | 47 | 0.32 | 4088 | 27.6 | 1530 |
| UNITED KINGDOM | 88 | 55 | 33 | 0.38 | 2857 | 32.5 | 1246 |
| GERMANY | 78 | 43 | 35 | 0.45 | 2797 | 35.9 | 924 |
| SPAIN | 65 | 45 | 20 | 0.31 | 2722 | 41.9 | 789 |
| IRAN | 56 | 42 | 14 | 0.25 | 2278 | 42.2 | 367 |
| JAPAN | 54 | 45 | 9 | 0.17 | 1728 | 30.9 | 757 |
| FRANCE | 52 | 35 | 17 | 0.33 | 1604 | 30.8 | 612 |
| AUSTRALIA | 40 | 33 | 7 | 0.18 | 1575 | 39.4 | 395 |

**Table 3.** The top 10 most productive institutions related to Epigenetic modifications in autoimmune diseases.

| Affiliation | Articles | Country | Citation | TLS |
| --- | --- | --- | --- | --- |
| CENT SOUTH UNIV. | 86 | China | 2880 | 1598 |
| UNIV. MICHIGAN | 84 | USA | 2340 | 1056 |
| TABRIZ UNIV. MED SCI | 81 | Iran | 385 | 134 |
| UNIV. CALIF DAVIS | 65 | USA | 2813 | 1344 |
| UNIV. TEHRAN MED SCI | 65 | Iran | 1366 | 240 |
| UNIV. PENN | 64 | USA | 2157 | 219 |
| HARV.ARD MED SCH | 47 | USA | 2079 | 340 |
| ANHUI MED UNIV. | 46 | China | 599 | 176 |
| FUDAN UNIV. | 45 | China | 688 | 353 |
| UNIV. CALIF SAN FRANCISCO | 45 | USA | 2598 | 394 |

### Analysis of Main Publishing Institutions and Authors

This study involves 2175 institutions that have published articles on epigenetic modifications in autoimmune diseases from 2012 to 2022. Table 2 presents the top ten institutions with the highest publication volume, with the United States, China, and Iran having the highest number of institutions. The top five institutions are Central South University (86 articles, 4.60%), University of Michigan (84 articles, 4.49%), Tabriz University of Medical Sciences (81 articles, 4.33%), University of California, Davis (65 articles, 3.48%), and Tehran University of Medical Sciences (65 articles, 3.48%). Central South University has the highest number of citations, reaching 2880. Additionally, we utilized the VOS viewer to create a visualization of the collaborative network among institutions (including only those with five or more publications). The network diagram consists of 16 nodes and 940 edges, as shown in Figure 4A. The top three institutions with the strongest collaboration capabilities are Central South University (TLS=1598), University of California, Davis (TLS=1344), and University of Michigan (TLS=1056). Central South University maintains close collaboration with institutions from the United States, China, and other countries.

This study involved a total of 9163 authors and 75956 co-cited authors. Tables 4A and 4B list the top ten authors in terms of publication volume and citation count, respectively. The H-index is a measure of a scholar’s overall influence, representing the number of publications (N) that have been cited at least N times. From Table 4A and Figure 4B, we can observe that Lu QJ and Zhao M have the highest H-indices, with values of 33 and 26, respectively, indicating their significant influence in the field. Furthermore, "Articles Fractionalized" is an indicator of an author’s contribution to a specific field, representing the proportion of an author’s publications in that field out of all publications.Lu QJ and Chang C are the top two contributors in this field, with values of 13.54 and 11.78, respectively. Additionally, Figure 4C displays a visualization of the collaborative network among authors using VOSviewer. Nodes represent authors, edges represent collaboration relationships, node size reflects the number of citations, and edge thickness represents the strength of collaboration. The top three authors with the highest citation counts are Lu QJ (2859 citations), Gershwin ME (1828 citations), and Zhao M (1668 citations), while the top three authors with the highest publication volume are Lu QJ (94 articles), Gershwin ME (88 articles), and Zhao M (87 articles). Furthermore, co-cited authors are authors who are simultaneously cited in one or more works. As shown in Table 4B, the top ten co-cited authors have each been cited over 100 times. Among them, the top five authors with the highest citation counts are Zhao M (315 citations), Hedrich CM (260 citations), Selmi C (264 citations), Lu QJ (201 citations), and Coit P (222 citations). Moreover, Figure 4D presents a visualization of the network among co-cited authors using VOSviewer, where the five authors with the highest total link strength are Zhao M (TLS=15847), Hedrich CM (TLS=12255), Selmi C (TLS=11136), Lu QJ (TLS=8937), and Coit P (TLS=8780).

**Table 4A.**
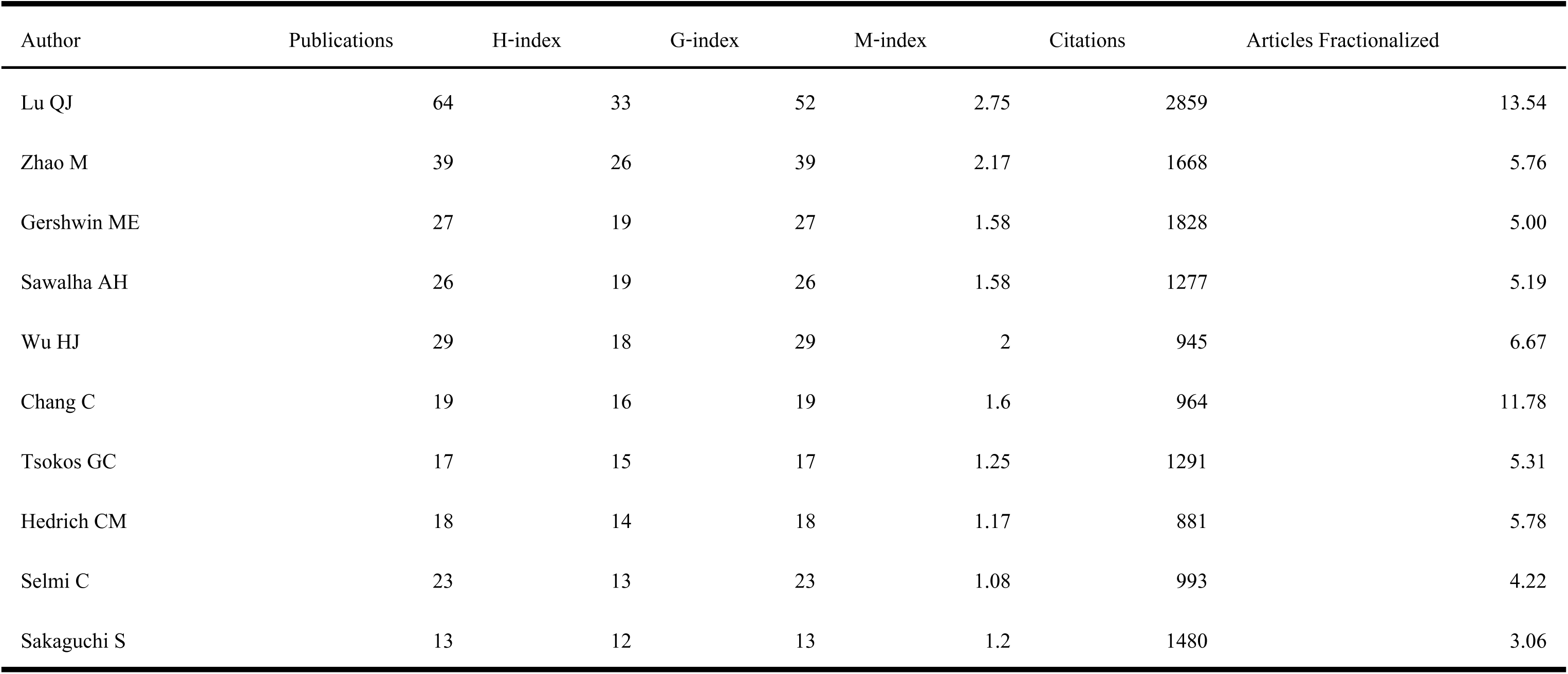
The top 10 most productive authors related to Epigenetic modifications in autoimmune diseases.

**Table 4B.**
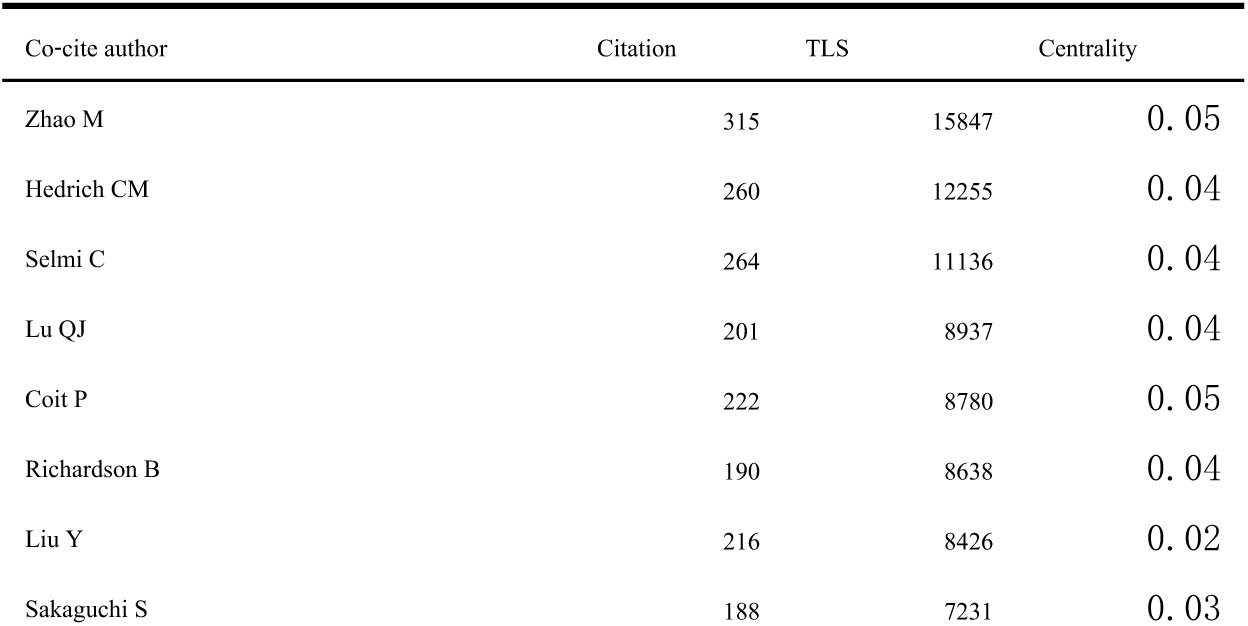
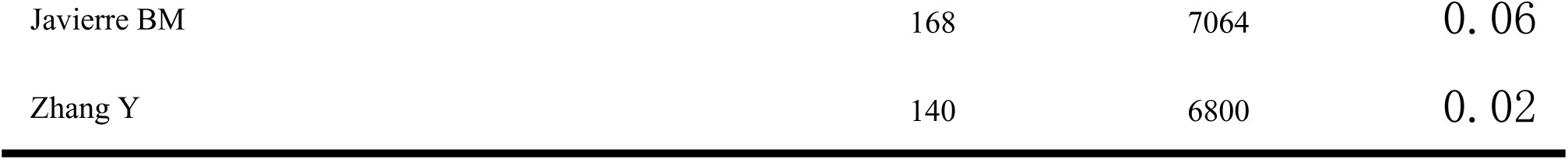
The top 10 most productive Co-cite author related to Epigenetic modifications in autoimmune diseases.

### Distribution of Source Journals and the Top 10 Highly Cited Articles

Research articles related to epigenetic modifications in autoimmune diseases have been published in 643 different journals, with 32 journals having published at least 10 articles. Table 5 presents the top 10 journals with the highest number of publications, accounting for 440 articles, which represents 23.53% of the total. Among the journals with more than 100 articles, Frontiers in Immunology (n=128) has the highest number, followed by Journal of Autoimmunity (n=71), International Journal of Molecular Sciences (n=56), Autoimmunity Reviews (n=35), and Clinical Immunology (n=33). Additionally, AM J HUM GENET (n=91) and AM J PATHOL (n=87) are also prominent journals in this field. The journal with the highest academic impact in this field is Journal of Autoimmunity, with an H-index of 45 and an impact factor of 14.51. Furthermore, Table 5 indicates that Journal of Autoimmunity (n=4530) is the most highly cited among the top 10 journals, followed by Frontiers in Immunology (n=2734).

**Table 5.** Top10 journals related to the research of Epigenetic modifications in autoimmune diseases.

| Rank | Journal | Publications | H-index | Total Citations | IF(2022) |
| --- | --- | --- | --- | --- | --- |
| 1 | Frontiers in immunology | 128 | 30 | 2734 | 8.79 |
| 2 | Journal of autoimmunity | 71 | 45 | 4530 | 14.51 |
| 3 | International Journal of Molecular Sciences | 56 | 18 | 872 | 6.21 |
| 4 | Autoimmunity Reviews | 35 | 23 | 1788 | 17.39 |
| 5 | Clinical immunology | 33 | 15 | 738 | 4.337 |
| 6 | Clinical reviews in allergy& immunology | 32 | 20 | 1065 | 10.82 |
| 7 | Proceedings of the national academy of sciences of the united states of America | 24 | 20 | 1596 | 12.78 |
| 8 | Current opinion in rheumatology | 22 | 17 | 838 | 4.94 |
| 9 | Nature communications | 22 | 16 | 1030 | 17.69 |
| 10 | Clinical Epigenetics | 17 | 15 | 628 | 7.26 |

**Table 6.**
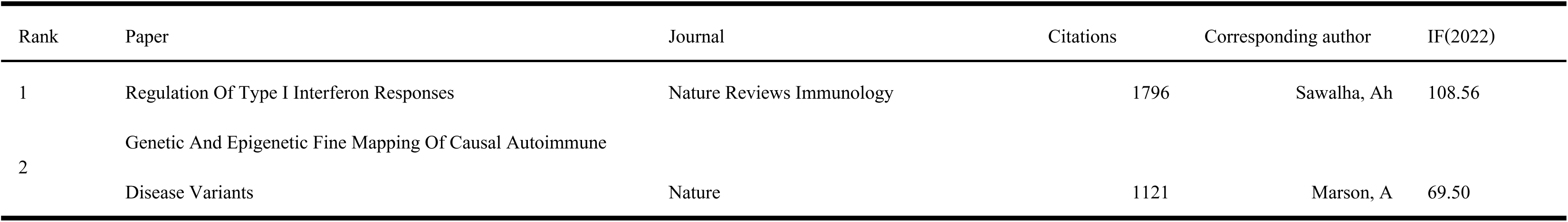
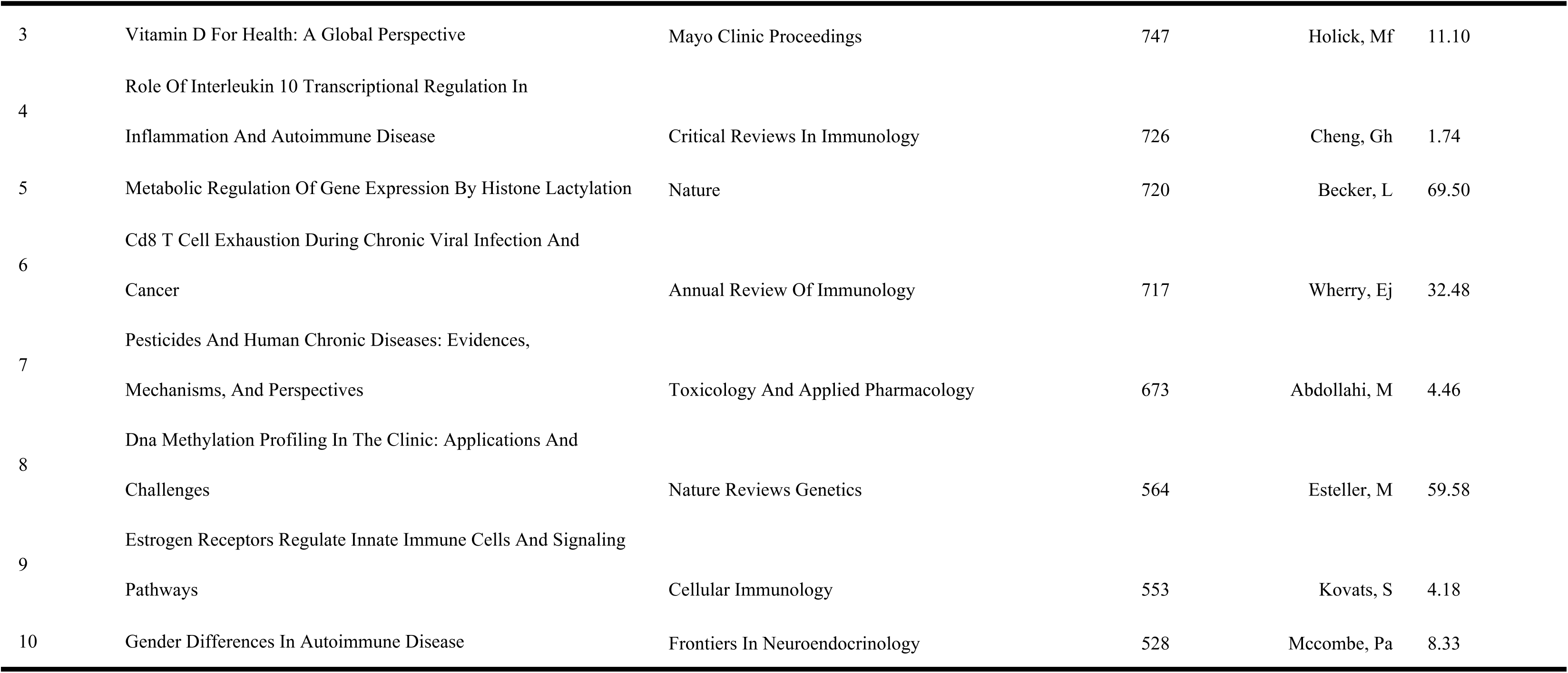
Top ten Aaticles with the most citations concerning the research of Epigenetic modifications in autoimmune diseases.

Moreover, we analyzed the annual publication trends of the top five most productive journals, as shown in Figure 5B. Frontiers in Immunology exhibits the highest growth rate, particularly in the past two years, followed by the International Journal of Molecular Sciences. We also conducted a visualization analysis of the popular research directions using R software, as depicted in Figure 5A. Currently, the top eight research directions in this field are immunology, biochemistry & molecular biology, cell biology, genetics & heredity, medicine, research & experimental, pharmacology & pharmacy, and endocrinology & metabolism.

Next, we present the top ten most cited articles in the field of epigenetic modifications in autoimmune diseases. The article titled "Regulation of Type I Interferon Responses" by Sawalha, Ah et al. has the highest citation count (1796 citations) and an impact factor of 108.56 (2022). Following that is the article by Marson, A et al. published in Nature, titled "Genetic and Epigenetic Fine Mapping of Causal Autoimmune Disease Variants," which has been cited 1121 times and has an impact factor of 69.50 (2022).

Furthermore, we used Citespace to illustrate the distribution of these journals and their citing journals(Figure 5C). The journals that cite others are depicted on the left side of the map, while the cited journals appear on the right side. The two main citation paths, represented by green and orange lines, demonstrate that researchers publish their studies in molecular/biological/immunological journals and medical/clinical journals, with a majority of citations originating from articles published in molecular/biological/genetic journals.

### Reference Analysis

In this study, a total of 116,521 references were cited, covering research related to the epigenetic modifications of autoimmune diseases. We identified the top five most cited references, as shown in Table 7. Among them, the article titled "Changes in the Pattern of DNA Methylation Associate with Twin Discordance in Systemic Lupus Erythematosus" by Javierre BM et al., published in Genome Research, has been cited 121 times and is considered a classic in this field. Following that is the article by Lu Q et al., titled "Demethylation of CD40LG on the Inactive X in T Cells from Women with Lupus," published in the Journal of Immunology, which has been cited 105 times and reveals the relationship between X chromosome inactivation and autoimmune diseases.

**Table 7.** Top five cited references concerning the research of Epigenetic modifications in autoimmune diseases.

| Rank | Reference | Journal | Citations | Corresponding author | IF (2022) |
| --- | --- | --- | --- | --- | --- |
| 1 | Changes in the pattern of DNA methylation associate with twin discordance in systemic lupus erythematosus | Genome Research | 121 | JAVIERRE BM | 9.44 |
| 2 | Demethylation of CD40LG on the inactive X in T cells from women with lupus | Journal of Immunology | 105 | LU Q | 5.43 |
| 3 | Genome-wide DNA methylation study suggests epigenetic accessibility and transcriptional poising of interferon-regulated genes in naïve CD4+T cells from lupus patients | Journal of Autoimmunity | 93 | COIT P | 14.51 |
| 4 | Genetic and epigenetic fine mapping of causal autoimmune disease variants | Nature | 87 | FARH KKH | 69.50 |
| 5 | Evidence for impaired t cell dna methylation in systemic lupus erythematosus and rheumatoid arthritis | ARTHRITIS RHEUM | 87 | RICHARDSO B | 15.483 |

Additionally, we employed VOSviewer to visualize the co-citation network of the most commonly cited references, as depicted in Figure 6A. This visualization illustrates the degree of association among the most cited references. Furthermore, we used Citespace to generate a burst detection visualization of reference citations, as shown in Figure 6B, highlighting the top 25 references with the strongest burst intensity. Notably, a review article titled "The Emerging Role of Epigenetics in Human Autoimmune Disorders" by Antonello Mai et al., published in Clinical Epigenetics, had the highest burst intensity in 2019, reaching 15.6. This indicates that the article has become one of the emerging hot topics in this field. The article by Javierre BM et al. had a burst intensity of 15.39, reaching its citation peak during the period of 2010-2015.

### Analysis of Keywords and Subject Terms

By analyzing the keywords in the abstracts and titles of publications, we can gain insights into the research interests and directions in the field. We collected a total of 5,278 subject terms, out of which 614 met the threshold criteria. Table 8 and Figure 7A present the top 5 subject terms with the highest citation counts, namely "expression"(329,7%),"DNA methylation"(295, 6%),"gene-expression"(225, 5%), "systemic-lupus-erythematosus" (220, 5%), and "rheumatoid arthritis" (204, 4%).The three diagrams represent a method of visually displaying the strength of connections between three main observation elements, such as authors, keywords, and institutions. Figure 7B illustrates the connections between authors, keywords, and institutions. The most frequently used keywords are "systemic-lupus-erythematosus," "DNA methylation," "expression," "gene-expression," and "rheumatoid-arthritis." Authors LU QJ, SELMI C., ZHAO M, and GERSHWIN ME have strong associations with the keywords "systemic-lupus-erythematosus," "expression," "DNA methylation," and "rheumatoid arthritis." Additionally,it can be observed that the majority of articles related to the keywords "systemic-lupus-erythematosus," "DNA methylation," and "expression" were published in Frontiers in immunology and Journal of autoimmunty. Frontiers in immunology and Clinical immunology encompass the main keywords shown in the diagram.

**Table 8.** Top ten Keywords with the most Frequences concerning the research of Epigenetic modifications in autoimmune diseases.

| Rank | Keywords | Frequency | TLS | Rank | Keywords | Frequency | TLS |
| --- | --- | --- | --- | --- | --- | --- | --- |
| 1 | expression | 329 | 1879 | 11 | differentiation | 131 | 827 |
| 2 | dna methylation | 295 | 1677 | 12 | cells | 102 | 561 |
| 3 | gene-expression | 225 | 1278 | 13 | association | 99 | 556 |
| 4 | systemic-lupus-erythematosus | 220 | 1297 | 14 | multiple-sclerosis | 93 | 540 |
| 5 | rheumatoid-arthritis | 204 | 1254 | 15 | autoimmunity | 92 | 561 |
| 6 | t-cells | 201 | 1167 | 16 | gene | 88 | 523 |
| 7 | regulatory t-cells | 150 | 877 | 17 | nf-kappa-b | 85 | 401 |
| 8 | genome-wide association | 143 | 799 | 18 | autoimmune-diseases | 84 | 434 |
| 9 | activation | 141 | 850 | 19 | risk | 83 | 442 |
| 10 | disease | 135 | 739 | 20 | pathogenesis | 78 | 459 |

Systemic lupus erythematosus(SLE) and rheumatoid arthritis(RA), being two types of autoimmune diseases, hold relevance in the study of autoimmune diseases. We conducted cluster analysis of keywords using VOSviewer, with a minimum co-occurrence frequency set at 5. Figure 7C demonstrates how keywords are grouped into ten clusters, each represented by a different color (red, pink, light red, green, dark blue, light blue, purple, brown, orange-yellow, and orange). Hypomethylation, expression, and gene expression are among the frequently used terms.

Furthermore, Figure 7D presents a timeline view of the keywords, depicting how research interests have evolved over time. The figure displays nine clusters, with "regulatory T cell" (#1) being the largest and consistently attracting high attention. "cAMP-responsive element modulator alpha" (#0), "cell-specific enhancer" (#6), and "genetic susceptibility" (#3) are current research hotspots. Figure 7E represents the burst strength of subject terms, which also reflects research frontiers, hot topics, and emerging trends. For example, "expression" was the most cited keyword in 2019. Recent popular topics such as "mroRNA" (2019-2022), "subsets" (2020-2022), "annotation" (2020-2022), and "diagnosis" (2016-2022) have bursts that extend until 2022.

Lastly, we analyzed the trend of subject evolution from 2012 to 2022, with 2018 as the time node, as shown in Figure 7F. "Differentiation" evolved into "activation," "expression" branched into "expression" and "activation," "DNA methylation" evolved into "expression," "gene expression," and "activation," "genome-wide association" evolved into "expression" and "gene expression," and "systemic-lupus-erythematosus" evolved into "gene expression."

## Discussion

### Global Overview of Epigenetic Modifications in Autoimmune Disease Research

This article presents a bibliometric analysis of epigenetic modifications related to autoimmune diseases between 2012 and 2022, aiming to understand the development trends and key topics in this field. Figure 1A demonstrates a continuous growth in the number of publications in this field over the past decade, particularly showing a rapid increase since 2018. This reflects the increasing attention and possibly signifies the entry into a period of rapid development in this field. The United States, China, and Italy are the top three countries in terms of publication volume and citation count. The United States ranks first in terms of publication volume, citation count, and average citations per article, indicating its dominant position in this field. China ranks second in terms of total citations and publication volume but has a lower average citation count. Italy ranks second in terms of average citation count with a value of 42.2. The network visualization graph of countries shows that the United States has the highest TLS (Total Link Strength), indicating its strong international influence. Another interesting finding is that while the United States has the greatest international influence, Germany has the highest MCP-Ratio, suggesting close collaboration between German researchers and researchers from other countries. In terms of research institutions, half of the top ten institutions with the highest publication output are from the United States, which explains why the United States contributes to a large number of articles related to epigenetic modifications and autoimmune diseases. This indicates that the establishment of high-level academic institutions plays an important role in academic research. Additionally, among the top ten institutions, China and Italy account for 30% and 20%, respectively. Among the top ten most productive authors, LU QJ has published the highest number of articles, reaching 64, and has the highest H-index with a citation count of 2859, demonstrating their highest academic influence in the field of epigenetics and autoimmune disease research and their leadership role in this field. Regarding journals, Table 4 lists journals such as Frontiers in Immunology, Journal of Autoimmunity, International Journal of Molecular Sciences, Autoimmunity Reviews, and Clinical Immunology, which are likely to be the main publishing venues for articles related to immunological diseases. It is recommended to submit more immunology-related articles to these journals. Among the most productive publications, five journals have an impact factor exceeding 10, namely Nature Communications (IF 2022, 17.69), Autoimmunity Reviews (IF 2022, 17.39), Journal of Autoimmunity (IF 2022, 14.51), Proceedings of the National Academy of Sciences of the United States of America (IF 2022, 12.78), and Clinical Reviews in Allergy & Immunology (IF 2022, 10.82). It is worth mentioning that these top 10 journals are excellent choices for researchers to publish high-quality research on epigenetic modifications and autoimmune diseases in the future. Furthermore, Figure 5A indicates that Immunology is the most popular research direction in this study. The impact of the top ten most cited articles is evaluated in Table 6. The most cited article is "Regulation Of Type I Interferon Responses," which may serve as a therapeutic target for autoimmune diseases such as SLE ^[23–25]^. It is worth noting that this article has a recent impact factor of 108.56, indicating its high quality. Marson, A et al., wrote a study titled "Genetic And Epigenetic Fine Mapping Of Causal Autoimmune Disease Variants," which focuses on the role of genetic and epigenetic fine mapping in autoimmune diseases ^[26]^. Similarly, this article also has a very high impact factor and is published in the top-tier journal Nature. Among these ten articles, the majority of the topics are concentrated on the relationship between autoimmune diseases, epigenetic modifications, etiology, diagnosis, and treatment. Furthermore, through co-citation analysis of the references, we can identify the current hot topics in this field as well as the researchers and institutions making outstanding contributions. As shown in Table 7, the article "Changes in the pattern of DNA methylation associate with twin discordance in SLE" by JAVIERRE BM has been cited 121 times, possibly making it the most frequently cited reference in this field. In Figure 7B, among the top 25 articles with the strongest citation bursts, there is a considerable number of studies related to epigenetic associations in SLE, indicating that SLE may be one of the representative autoimmune diseases associated with epigenetic modifications. Moreover, most of these references elucidate the pathophysiology, diagnosis, and treatment of autoimmune diseases at the genetic level, indicating that these directions are hot research topics in the field of epigenetic modifications associated with autoimmune diseases."

### Current research hotspots of epigenetic modification in autoimmune diseases

In our study, we utilized co-occurrence networks of keywords based on the titles/abstracts of all included publications. Combining Figure 7C and 7D, we identified seven current hot research topics: regulatory T cells (Tregs), rheumatoid arthritis, epigenetic regulation, cAMP-responsive element modulator alpha, cell-specific enhancer, genetic susceptibility, and SLE. These results are consistent with the research hotspots in the field of epigenetic modifications and autoimmune diseases, and the relationships among these hotspots are summarized in Figure 8.(1)Regulatory T cells (Tregs): As research on Tregs advances, we are increasingly recognizing the significance of epigenetic regulation in Treg function and the development of autoimmune diseases. It has been found that genetic abnormalities and environmental damage affecting the quantity or function of Tregs can lead to autoimmune diseases ^[27]^. Mutations or abnormalities in other genes expressed in immune cells that impact Treg development and function may also contribute to autoimmune/inflammatory diseases, referred to as "Tregopathies" ^[28]^. In SLE, DNA methylation defects in Tregs are closely associated with disease progression ^[29]^. Furthermore, identifying suitable targets to regulate Treg function is a current research focus. For instance, histone deacetylase 9 (HDAC9) has been found to play a crucial role in Treg cells, regulating their immunosuppressive function ^[30]^. Inhibiting the expression of HDAC6 has been shown to improve Treg cell function ^[31]^, suggesting that this inhibitor could serve as a potential target for modulating immune function. Gaining a deeper understanding of the pathogenesis of autoimmune diseases provides new insights and potential targets for developing novel therapeutic approaches.(2)Rheumatoid arthritis (RA) is characterized by the abnormal expression of various inflammatory factors, and several key research hotspots have been identified through co-linearity analysis in Figure 7C. One important factor is growth factor-beta (TGF-β), which plays a significant role in the development and progression of RA. TGF-β is involved in regulating key pathological processes such as inflammation, fibrosis, and joint destruction. Studies have shown that TGF-β has a dual role in joint destruction. On one hand, it can promote synoviocyte proliferation and neovascularization, thereby exacerbating joint damage^[32]^. On the other hand, it can regulate osteoclast differentiation and function ^[33]^, thus alleviating joint destruction. Therefore, utilizing the regulatory effects of TGF-β, it can be applied in cell therapy to promote joint tissue repair and regeneration. This cell therapy strategy may involve the use of modified cells or gene therapy to enhance the biological effects of TGF-β.In addition to its role in joint destruction, TGF-β plays an important role in immune regulation. It can promote the development and function of regulatory T cells (Treg cells) while suppressing the activation and function of inflammatory T cells ^[34]^. By utilizing the immunoregulatory function of TGF-β, the activation and function of inflammatory T cells can be regulated, thereby alleviating the inflammatory response in the joints. This may involve using TGF-β or its related molecules as immunomodulators for RA treatment.Furthermore, recent studies on differential gene expression analysis have provided important insights into the pathophysiological processes of RA and the development of new therapeutic strategies. RA involves the dysregulated expression of various inflammatory factors. For example, studies have shown a significant increase in the expression of inflammatory cytokines such as IL-6, IL-17, and IL-23 in RA patients. Inhibiting the expression of these inflammatory factors or their signaling pathways may help alleviate inflammation and joint destruction in RA ^[35–37]^. Additionally, research has indicated that compared to healthy individuals, RA patients exhibit upregulated expression of genes in synoviocytes and osteoblasts, such as matrix metalloproteinases (MMPs) and receptor activator of NF-κB ligand (RANKL). Inhibiting the expression of these genes or their signaling pathways may contribute to slowing down the progression of joint damage ^[38–40]^.In summary, studies focusing on differential gene expression provide important clues for a deeper understanding of the pathogenesis and pathological processes of RA. These studies reveal aberrant expression of multiple key genes, pathways, and transcription factors in RA, offering potential targets for the development of new treatment strategies and drugs.

(3)Epigenetic modifications play a significant role in the occurrence and development of autoimmune diseases. Abnormal epigenetic modifications have been closely associated with the pathogenesis of autoimmune diseases, and some have shown promise as diagnostic and disease activity markers. For example, the hypomethylation of the IFI44L promoter is used for diagnosing SLE. Recently discovered non-coding RNAs, including long non-coding RNAs (lncRNAs) and circular RNAs (circRNAs), are also implicated in the etiology of autoimmune diseases. ^[41]^Differential DNA methylation levels of specific genes related to inflammation, joint destruction, and immune regulation have been observed in RA patients’ joint tissues. ^[42,43]^Furthermore, several validated lncRNAs, such as HOTAIR, GAS5, and HIX003209, have shown potential as novel biomarkers for diagnosis and treatment in RA.^[44]^(4)cAMP-responsive element modulator alpha (CREMα): CREMα is a transcription factor involved in the regulation of immune cell development, function, and stability. Recent studies have demonstrated that CREMα plays a critical role in autoimmune diseases and is associated with therapeutic interventions.^[45–47]^ Increased expression of CREMα in T cells of SLE patients promotes inflammatory cell activation and cytokine production, triggering arthritis inflammation.^[48]^ Moreover, CREMα is involved in regulating the function of regulatory T cells (Tregs). Overexpression of CREMα inhibits Treg cell suppressive functions, disrupting immune tolerance and enhancing autoimmune responses. ^[49–51]^Further research on the functions and regulatory mechanisms of CREMα in autoimmune diseases, as well as its associations with therapeutic interventions, may provide a theoretical basis for the development of more effective treatment strategies. (5)Cell-specific enhancers: Cell-specific enhancers play a crucial role in the epigenetic regulation. These enhancers are capable of modulating gene expression in specific cell types, thereby influencing the function of immune cells and the regulation of inflammatory responses ^[52–54]^. Research has found the presence of a type of cell-specific enhancer, such as rs2431697, in SLE patients. It may disrupt regulatory elements and regulate the expression of miR-146a, leading to SLE development ^[55]^. Additionally, the activation of intestinal epithelial cell-specific enhancers has been associated with impaired gut barrier function and enhanced inflammation in patients with inflammatory bowel disease (IBD) ^[56]^. These studies suggest that abnormal activation of cell-specific enhancers may play a significant regulatory role in the pathogenesis of autoimmune diseases. Further research will contribute to a deeper understanding of the role of epigenetic regulation in autoimmune diseases, providing new insights and targets for diagnosis and treatment.(6) Genetic susceptibility:Genetic susceptibility is closely related to the dysregulation of epigenetic regulation and the development of autoimmune diseases.

Studies have revealed differences in DNA methylation patterns between RA (Rheumatoid Arthritis) patients and healthy individuals. For instance, reduced DNA methylation levels of the immune-related gene TNFAIP3 were found in RA patients, leading to increased gene expression and subsequent promotion of inflammatory responses ^[57]^. Furthermore, the DNA methylation status of the HLA-DRB1 gene region was found to be closely associated with the occurrence and progression of RA ^[58]^. SLE, being a complex autoimmune disease, is also closely linked to genetic susceptibility genes and abnormal histone modifications. For example, certain genetic susceptibility genes in SLE patients are associated with patterns of histone acetylation and methylation, affecting immune cell function and the regulation of inflammatory responses ^[59]^. Another study found a correlation between genetic susceptibility genes in SLE patients and abnormal histone ubiquitination, resulting in hyperactivation of immune cells and increased autoimmune reactions ^[60]^. These studies provide important clues for a better understanding of the pathogenesis of autoimmune diseases and offer new avenues for developing personalized treatment strategies and precision medicine. (7) Systemic Lupus Erythematosus:SLE is a keyword that frequently appears in the context of DNA methylation and diagnosis. DNA methylation patterns in SLE patients differ from those in healthy individuals. Changes in DNA methylation of certain genes have been associated with the pathogenesis of SLE ^[61–63]^. Therefore, analyzing DNA methylation sites in peripheral blood cells of SLE patients can potentially identify differences between patients and healthy individuals, providing potential biomarkers for early diagnosis of SLE. Additionally, specific DNA methylation sites can serve as predictive indicators for SLE treatment. By examining the DNA methylation status of specific genes in patients, treatment response and disease progression can be predicted, offering a basis for personalized treatment. In terms of treatment, DNA methyltransferases (DNMTs) are key enzymes involved in DNA methylation regulation. Research has shown that the use of DNA methyltransferase inhibitors, such as 5-azacytidine, can reduce DNA methylation levels in SLE patients and worsen the disease condition ^[64,65]^. Further research on the relationship between DNA methylation and SLE, as well as the development of new diagnostic and treatment strategies, is essential to improve patient outcomes.

In summary, although there are many types of autoimmune diseases, most of the current research focuses on rheumatoid arthritis and systemic lupus erythematosus. These two diseases are currently the focus of research. The current research hot topics, such as regulatory T cells, growth factor β, epigenetic regulation, genetic susceptibility and CREMα, researchers strive to explore the pathogenesis and mechanism of autoimmune diseases through these topics. These research directions will help to reveal the etiology of the disease, discover new therapeutic targets, and develop more effective treatment strategies, thereby improving the quality of life of patients and reducing the severity of the disease. However, further studies and experiments are needed to fully understand these mechanisms and to provide more specific and individualized approaches for the treatment of autoimmune diseases.

### Research frontiers and future trends of epigenetic modifications in autoimmune diseases

The frontiers and future trends in research on epigenetic modifications in autoimmune diseases are reflected in the topic evolution graph, as shown in Figure 7F. The research direction has evolved from keywords like "differentiation," "expression," "DNA methylation," "genome-wide association," and "systemic lupus erythematosus" to "expression," "activation," and "gene expression," which significantly influence future researchers. As illustrated in Figure 7A, "expression" and "gene expression" are also the most frequently cited keywords, indicating their central position in autoimmune disease and epigenetic research. Aberrant regulation of gene expression is a crucial factor in the development of autoimmune diseases, and epigenetic mechanisms play a critical role in gene expression regulation ^[66]^. In RA, DNA methylation changes in immune-related genes such as HLA-DRB1 and PTPN22 are closely associated with disease risk and clinical manifestations ^[67,68]^. Moreover, abnormal regulation of gene methylation is also associated with the development and pathological features of autoimmune diseases such as systemic lupus erythematosus and Sjögren’s syndrome ^[69,70]^. Additionally, the abnormal expression of certain microRNAs is linked to the occurrence and progression of autoimmune diseases ^[71]^. Studies have found associations between single nucleotide polymorphisms (SNPs) and epigenetic markers in genetic susceptibility to autoimmune diseases ^[72,73]^. Future research should further explore the complexity of epigenetic regulatory networks and their specific roles in different types of autoimmune diseases. This will provide new opportunities for developing personalized treatment strategies and innovative immunotherapies.

Co-occurrence analysis of emerging terms reveals future research trends. Figure 7E highlights recent hot topics, namely "mroRNA" (2019-2022), "subsets" (2020-2022), and "annotation" (2020-2022), which are likely to continue receiving attention in the near future.Firstly, microRNA (miRNA) has garnered widespread interest as a significant regulatory factor in studying the epigenetic mechanisms of autoimmune diseases. MiRNAs are a class of small non-coding RNA molecules that regulate gene transcription and translation by binding to target gene mRNA. Increasing research focuses on novel biomarkers of miRNAs for the diagnosis and treatment of autoimmune diseases. Studies have found elevated expression levels of microRNA-21 and miR-155 in SLE patients, which are correlated with disease activity and clinical severity.^[74–75]^ MiR-155, through regulating the expression of multiple immune-related genes, influences the differentiation, function, and inflammatory response of immune cells, thereby participating in the pathogenesis of autoimmune diseases. ^[76]^Therefore, inhibiting the expression of microRNA-21 and microRNA-25 or using microRNA-21 and microRNA-25 antagonists may help alleviate inflammation and immune reactions in systemic lupus erythematosus. Currently, miRNA research is rapidly evolving, and further in-depth studies are needed to validate these potential mechanisms and explore the prospects of miRNA in the treatment of autoimmune diseases.

Next, the epigenetic regulation patterns of different immune cell subsets play a crucial role in the pathogenesis and progression of autoimmune diseases. Co-occurrence analysis indicates that "regulatory T cells," "Th17 cells," and "B cells" are the main research focuses. Th17 cells play an important role in the onset and progression of RA. ^[77]^Specific gene DNA methylation and histone modifications have been found to play a key role in the development and functional regulation of Th17 cells. For example, the DNA methylation level and histone acetylation level of the IL-17A gene are associated with disease activity in RA patients. ^[78–79]^These epigenetic markers can serve as indicators for the diagnosis and prognostic assessment of RA. B cells also play a significant role in the immunopathological processes of systemic lupus erythematosus (SLE). Studies have revealed correlations between specific gene DNA methylation and histone modification patterns in B cell subsets and disease occurrence and severity in SLE patients. ^[80–81]^Exploring the epigenetic regulatory mechanisms of cell subsets will contribute to uncovering the pathological processes of autoimmune diseases and provide new therapeutic targets and strategies for disease diagnosis, classification, and prognostic evaluation. Additionally, CAR-T cell therapy has made significant progress in cancer treatment and is beginning to be applied in the treatment of autoimmune diseases. Recently, Andreas Mackensen and colleagues achieved favorable results in treating refractory systemic lupus erythematosus with Anti-CD19 CAR T cell therapy. ^[82]^CAR-T cell therapy enables T cells to recognize and attack specific target cells by introducing a recombinant T cell receptor (CAR). It is believed that in the near future, there will be more precise and effective therapies for the treatment of autoimmune diseases.

Lastly, annotation-based research methods provide crucial clues for a deeper understanding of the association between epigenetic regulation and autoimmune diseases. By annotating specific genomic regions, we can uncover functional elements, modification states, and changes related to autoimmune diseases within those regions, thereby revealing the mechanisms of epigenetic regulation in disease onset and development. In recent years, annotation-based research methods have made significant progress in the field of autoimmune diseases. By annotating the composition and modification states of functional elements, researchers have discovered important regulatory regions for multiple autoimmune disease-related genes.^[83–85]^ These annotation insights not only contribute to understanding the functions and regulatory mechanisms of these genes but also provide new directions for further research. Moreover, annotation methods can reveal the association between changes in epigenetic modifications and the occurrence and development of autoimmune diseases, providing new avenues for early diagnosis and treatment of these diseases. Annotation-based research methods offer essential tools for a comprehensive understanding of the relationship between epigenetic regulation and autoimmune diseases. By annotating the composition and modification states of functional elements, we can reveal crucial regulatory regions and mechanisms, offering new clues for the occurrence and treatment of autoimmune diseases.

In conclusion,through extensive exploration of the association between epigenetic modification and autoimmune diseases, researchers have found that abnormal regulation of gene expression, gene methylation changes and abnormal expression of microrna are closely related to the occurrence and development of diseases. In addition, DNA methylation and histone modifications of specific genes play a key role in the regulation of immune cell subsets, such as Th17 cells and B cells. Future studies will further explore the complexity of the epigenetic regulatory network and its specific role in different types of autoimmune diseases, providing new opportunities for individualized treatment strategies and innovative immunotherapies. In addition, the latest research hotspots focus on microRNA, cell subset regulation, and annotation-based research methods, as shown in Figure 9. In-depth research in these fields is expected to reveal new mechanisms and provide new breakthroughs for the diagnosis and treatment of autoimmune diseases.

### Limitations

There are still some limitations that need to be discussed: (1) Due to limitations of the literature metrics software, not all studies collected from WoSCC, PubMed, Cochrane, Scopus, and Embase library databases were included, potentially leading to publication bias. Therefore, in future research, it is recommended to use more data sources and powerful software. (2) Due to the constant publication of new research every day, we may overlook some impactful new studies. (3) We only extracted research and review articles in English, excluding non-English publications or non-research/review articles, which may result in some omissions. (4) The data selection was done by two authors and discrepancies were resolved through consultation with experts to reach a final consensus.

### Conclusion

Through bibliometric analysis, we explored the relationship between epigenetic modifications and autoimmune diseases and analyzed the trends in international collaboration, authors, publications, keywords, and research hotspots. These findings are of significant importance in the medical field as they reveal new developments and trends in the field of epigenetic modifications in autoimmune diseases, advancing the study of epigenetics in autoimmune disease research. This study is the first comprehensive and systematic analysis of the global trends in epigenetic modifications related to autoimmune diseases over the past 11 years using bibliometric analysis. We provided a detailed overview of global publication trends, helping scholars identify key authors, institutions, and journals in the field. Furthermore, through keyword and co-citation cluster analysis, we provided guidance for researchers in selecting new research directions. These directions primarily focus on regulatory T cells (Tregs), rheumatoid arthritis, epigenetic regulation, cAMP-responsive element modulator alpha, cell-specific enhancer, genetic susceptibility, and systemic lupus erythematosus. Additionally, future research trends will involve elucidating the mechanisms of epigenetic regulation in disease occurrence and development through annotation of specific genomic regions, with a greater emphasis on targeting microRNAs through mimics or inhibitors and cell-based therapies for the treatment of autoimmune diseases. These approaches hold great value and promise in regulating various aspects of human autoimmune diseases. It can be predicted that further collaboration between authors, institutions, and countries in the future will accelerate the development of epigenetic research related to autoimmune diseases.

### Data Availability Statement

The original contributions presented in the study are included in the article/supplementary material. Further inquiries can be directed to the corresponding author.

### Author Contributions

Xiang Gao drafted the manuscript, XinHuang and Sheng Sun compiled the data, Tao Chen and Yehui Wang reviewed the manuscript, Yongxiang Gao and Xiaodan Zhang provided funding.

### Funding

This work was supported by the National Natural Science Foundation of China (82074329) ;Summary of experience in characteristic diagnosis and treatment of traditional Chinese medicine and inheritance and research of academic thought (No. 030054070);The inheritance and innovation Project of traditional Chinese Medicine (008080022);Inheritance Project of Academic thoughts of famous TCM doctors (003109011007).

## Data Availability

All relevant data are within the manuscript and its Supporting Information files.

N

## Acknowledgments

We would like to thank Web of Science Core Collection for providing the raw data for this study.

---

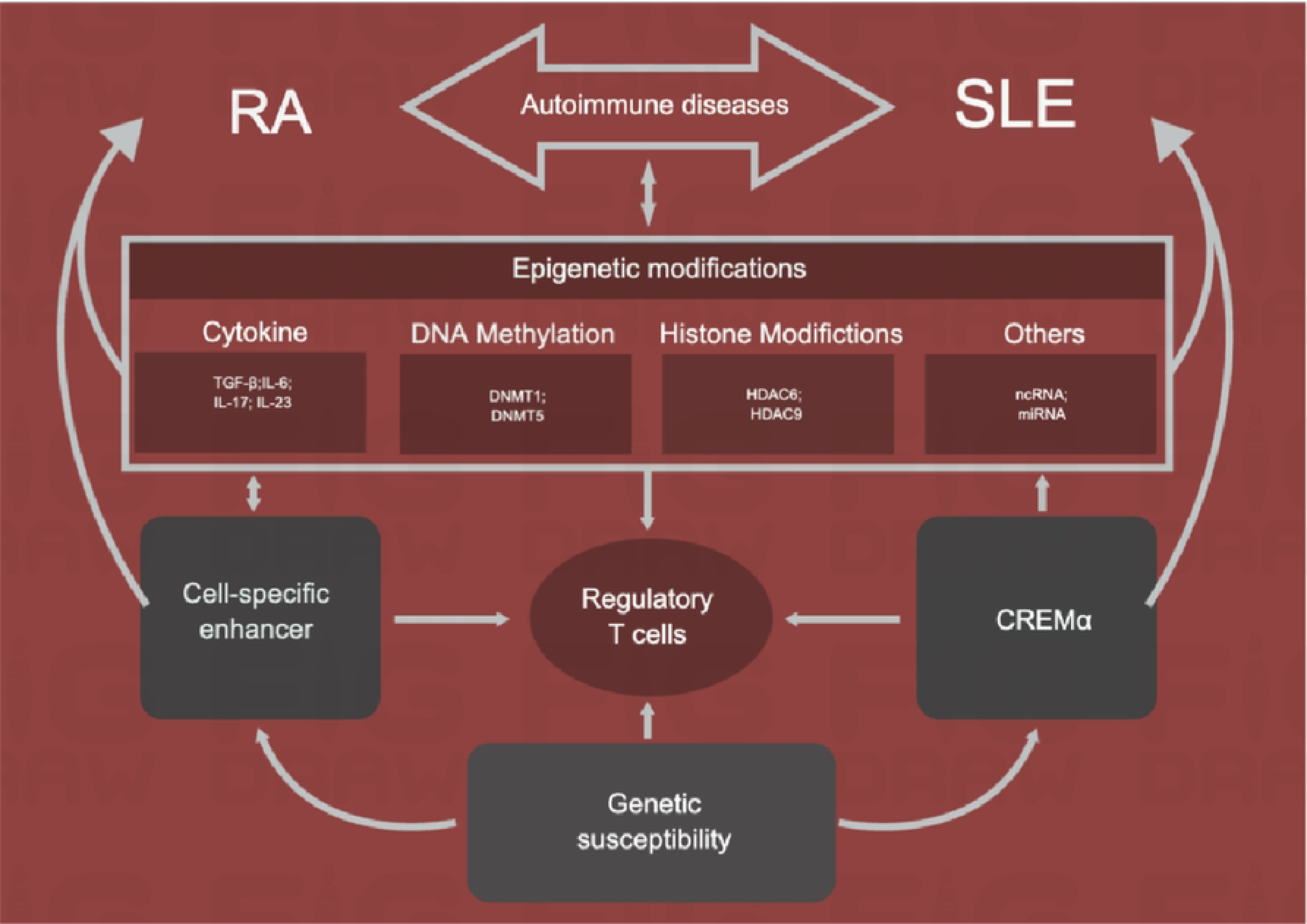

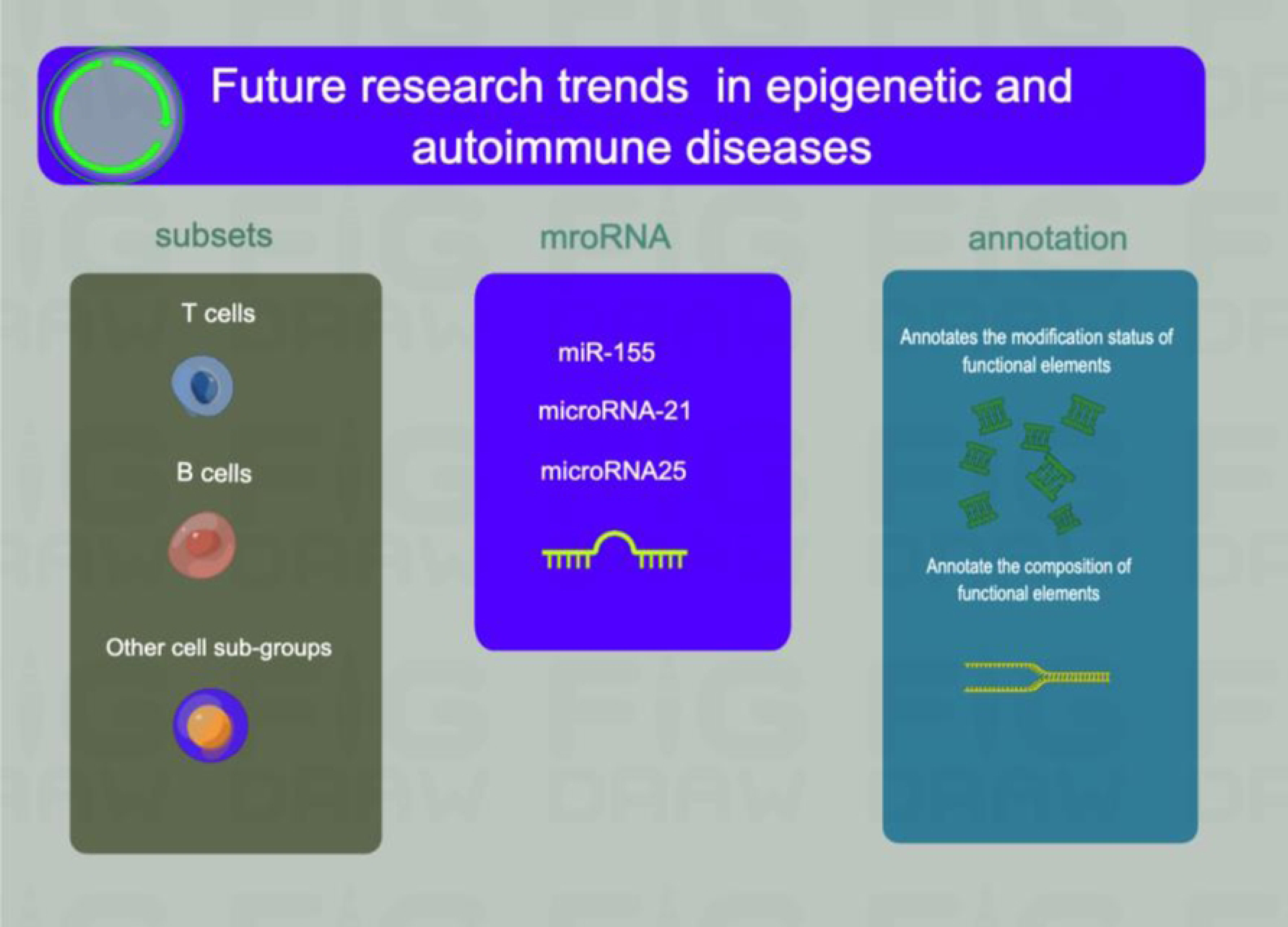

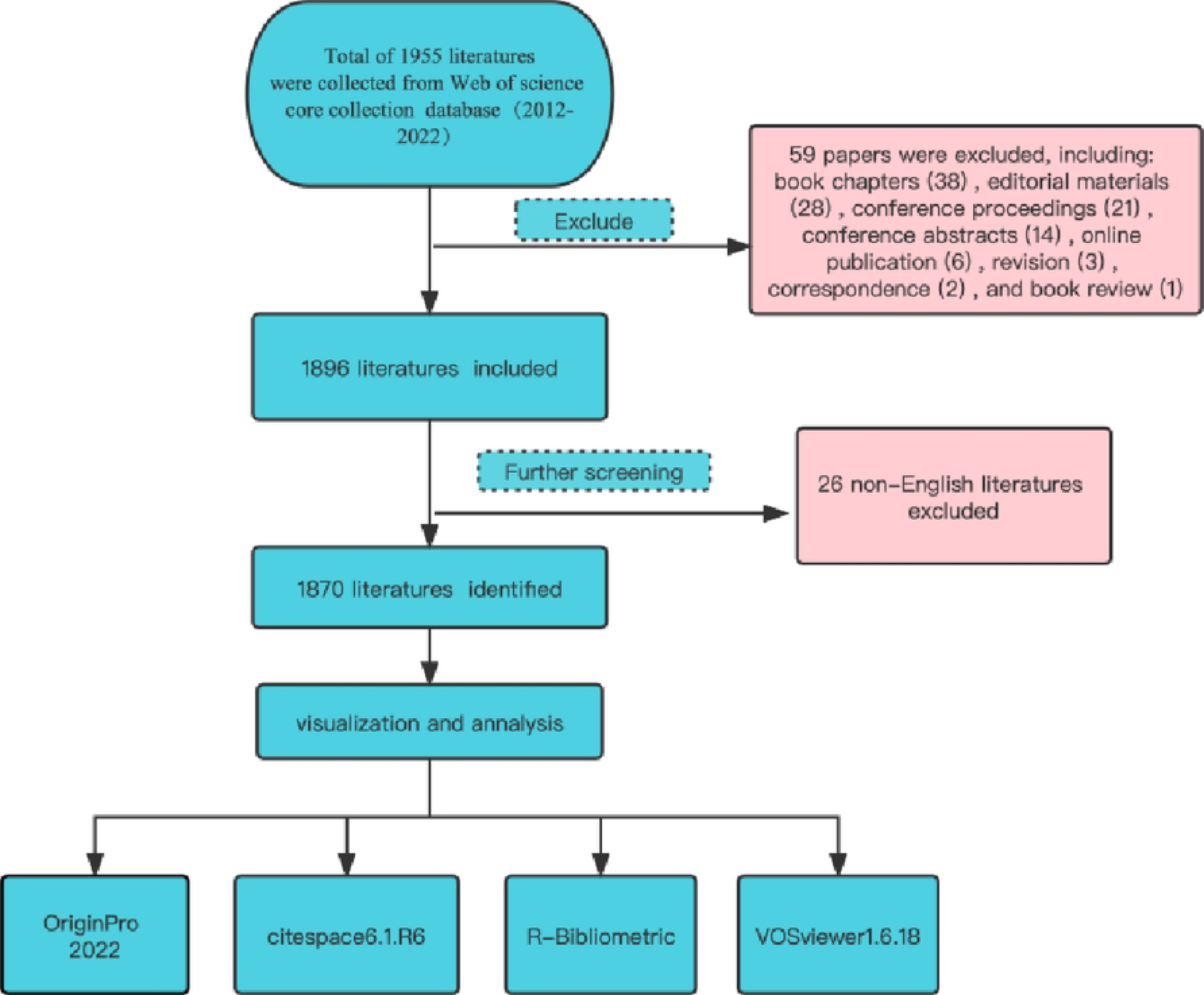

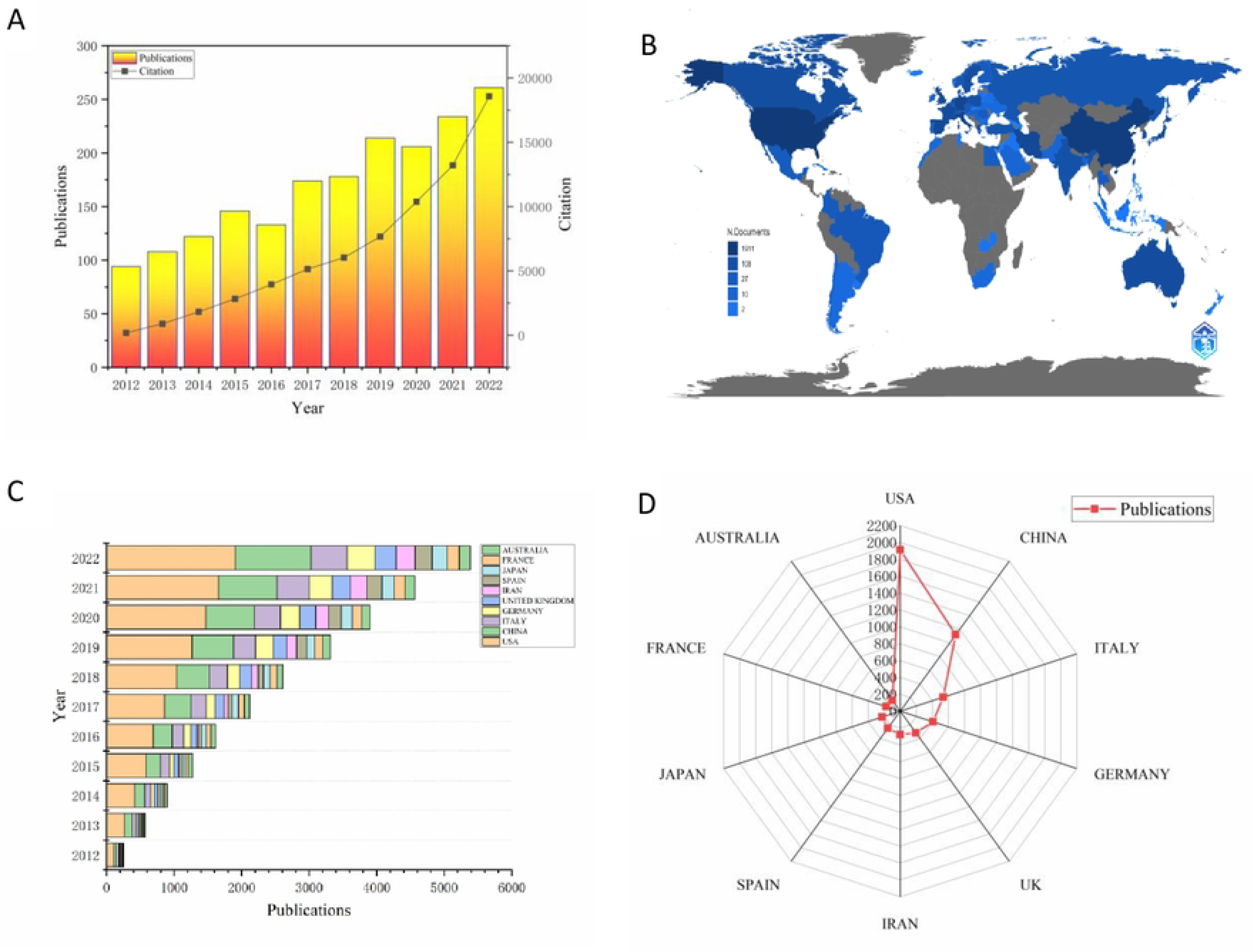

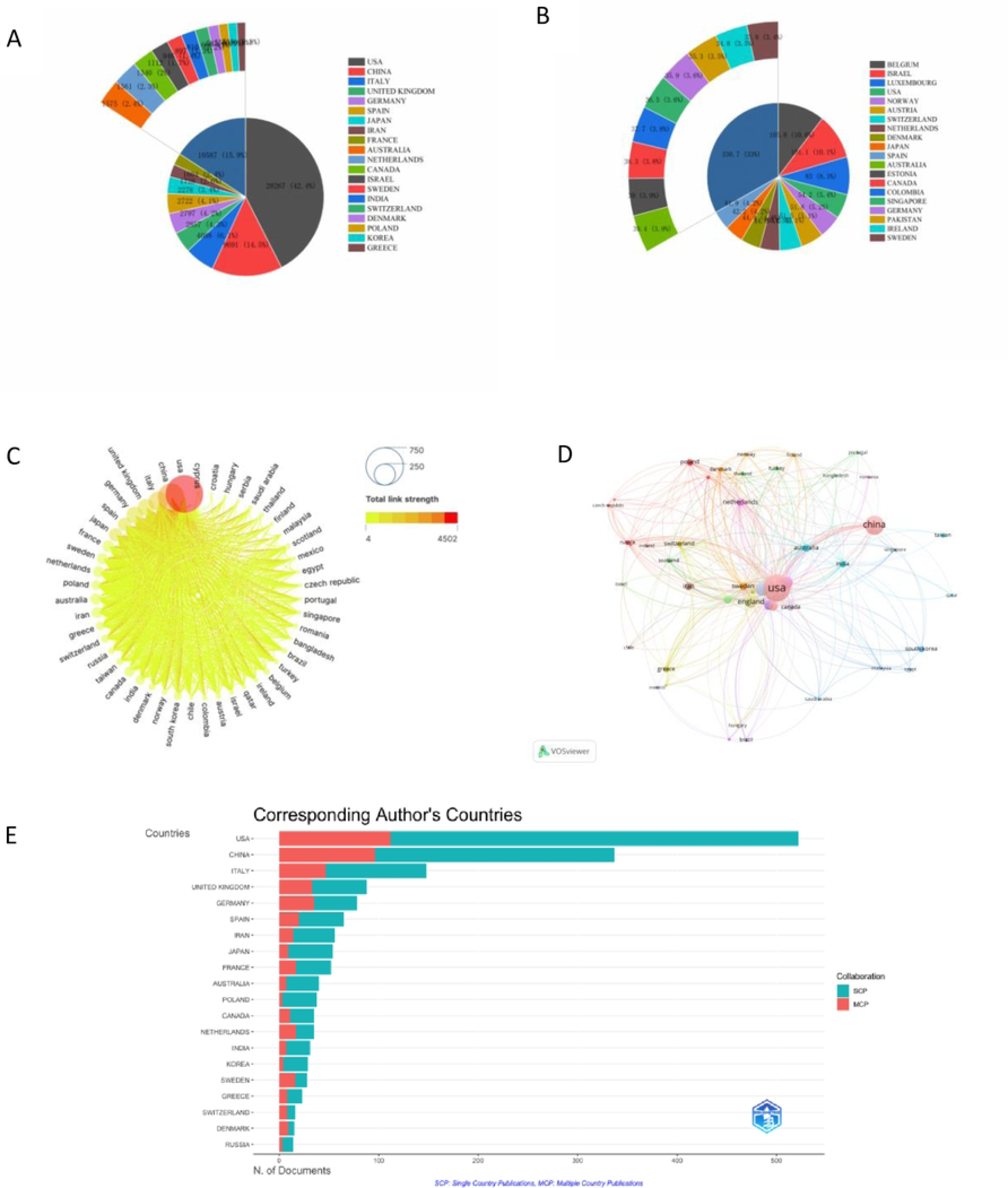

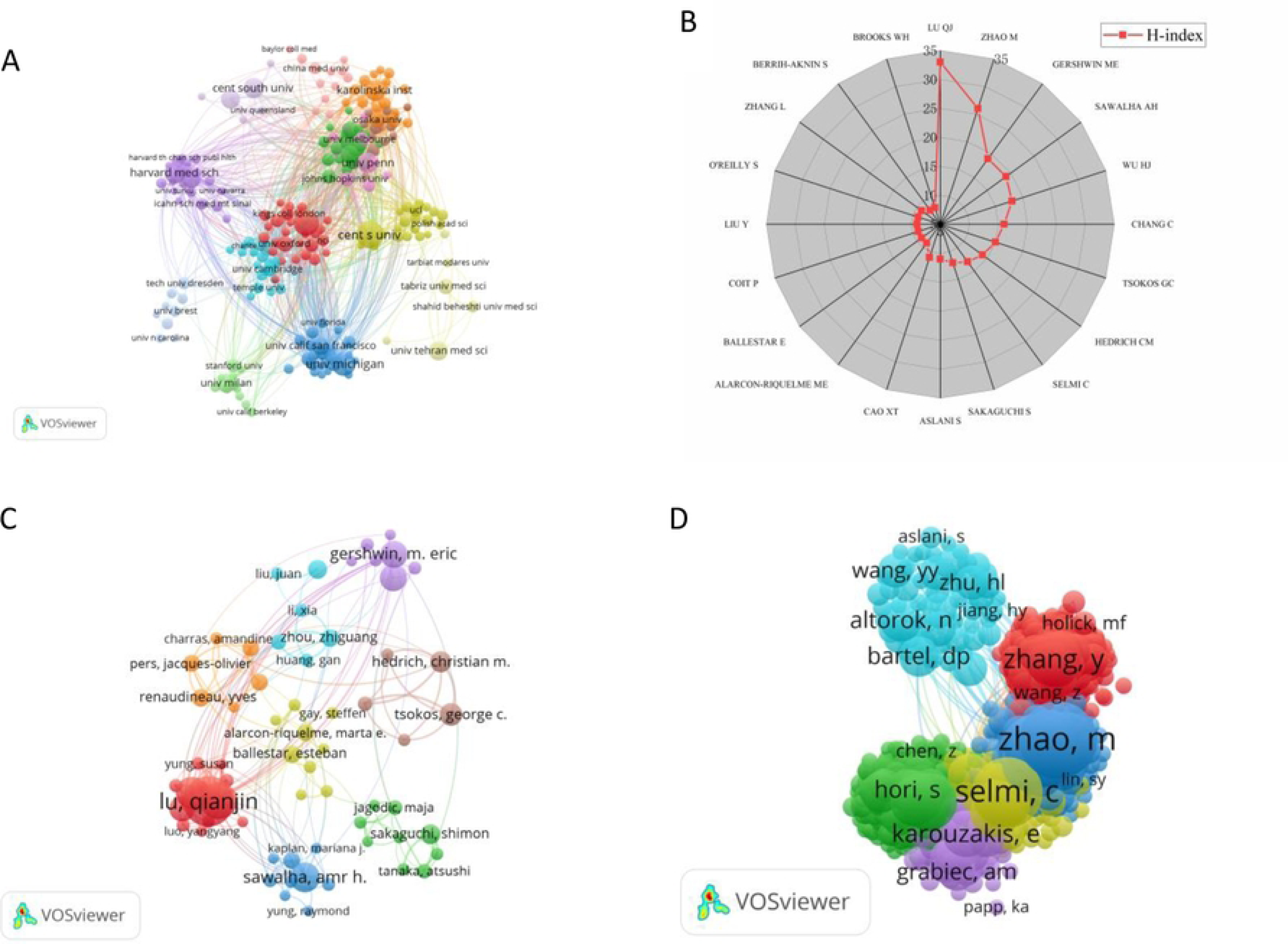

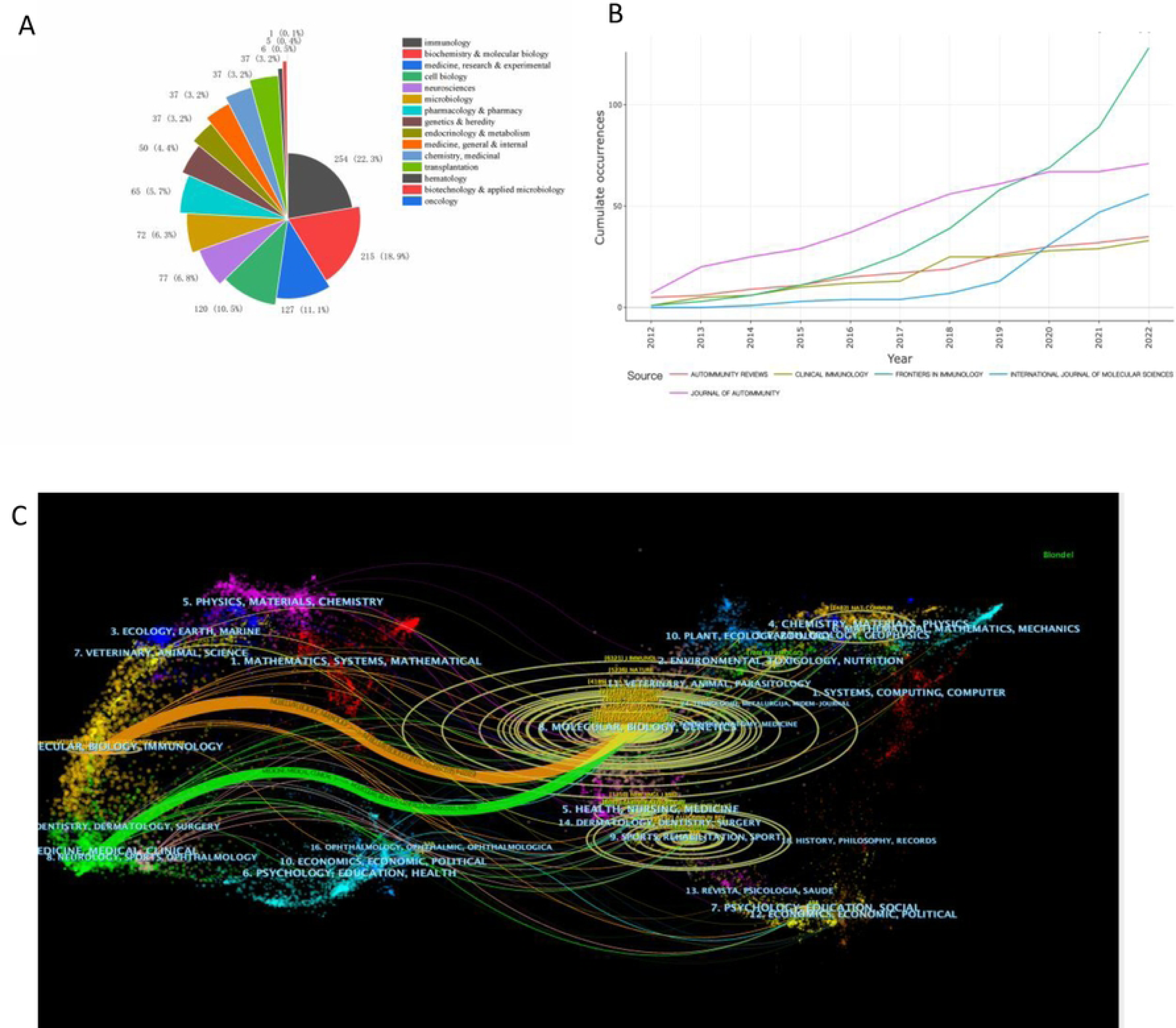

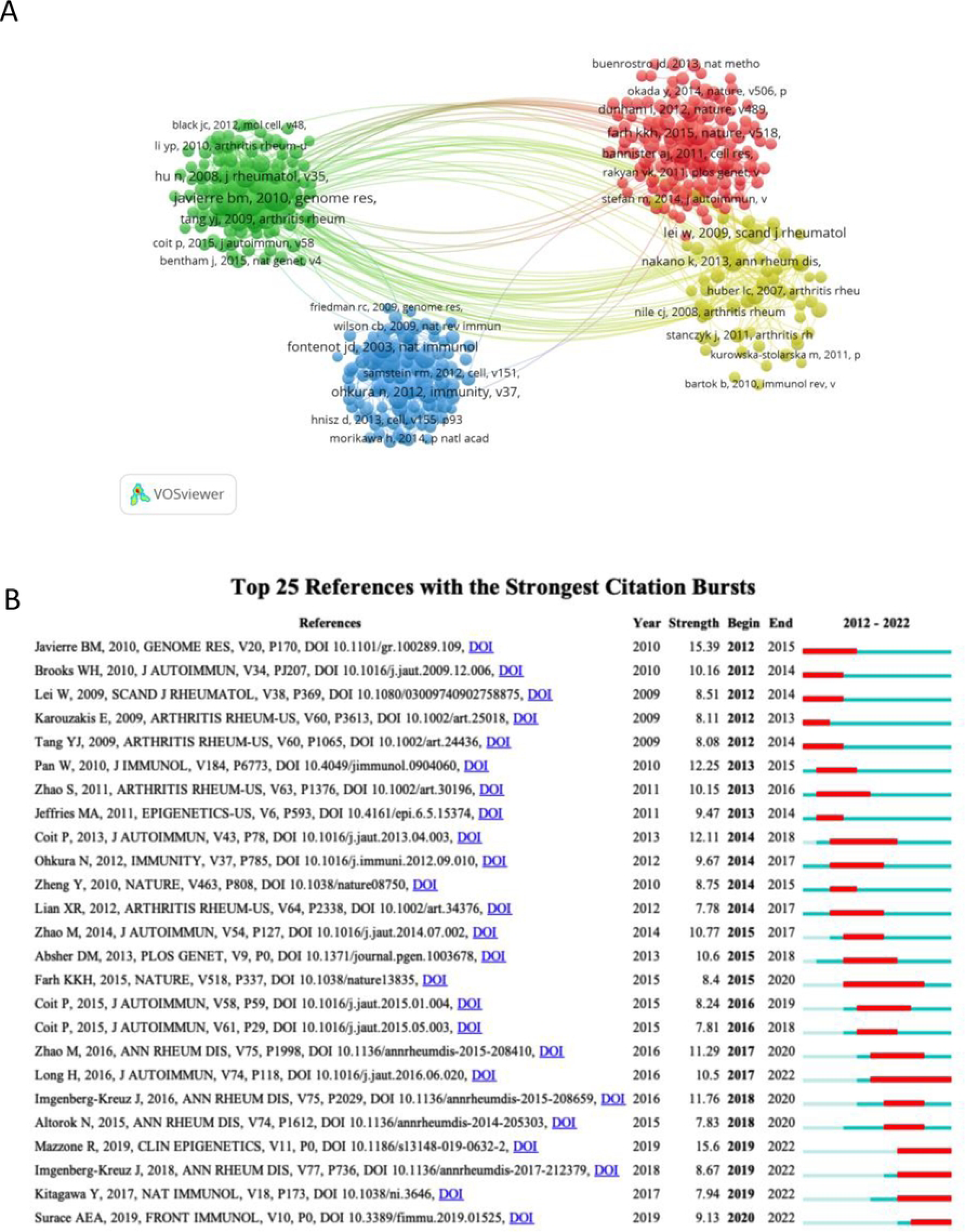

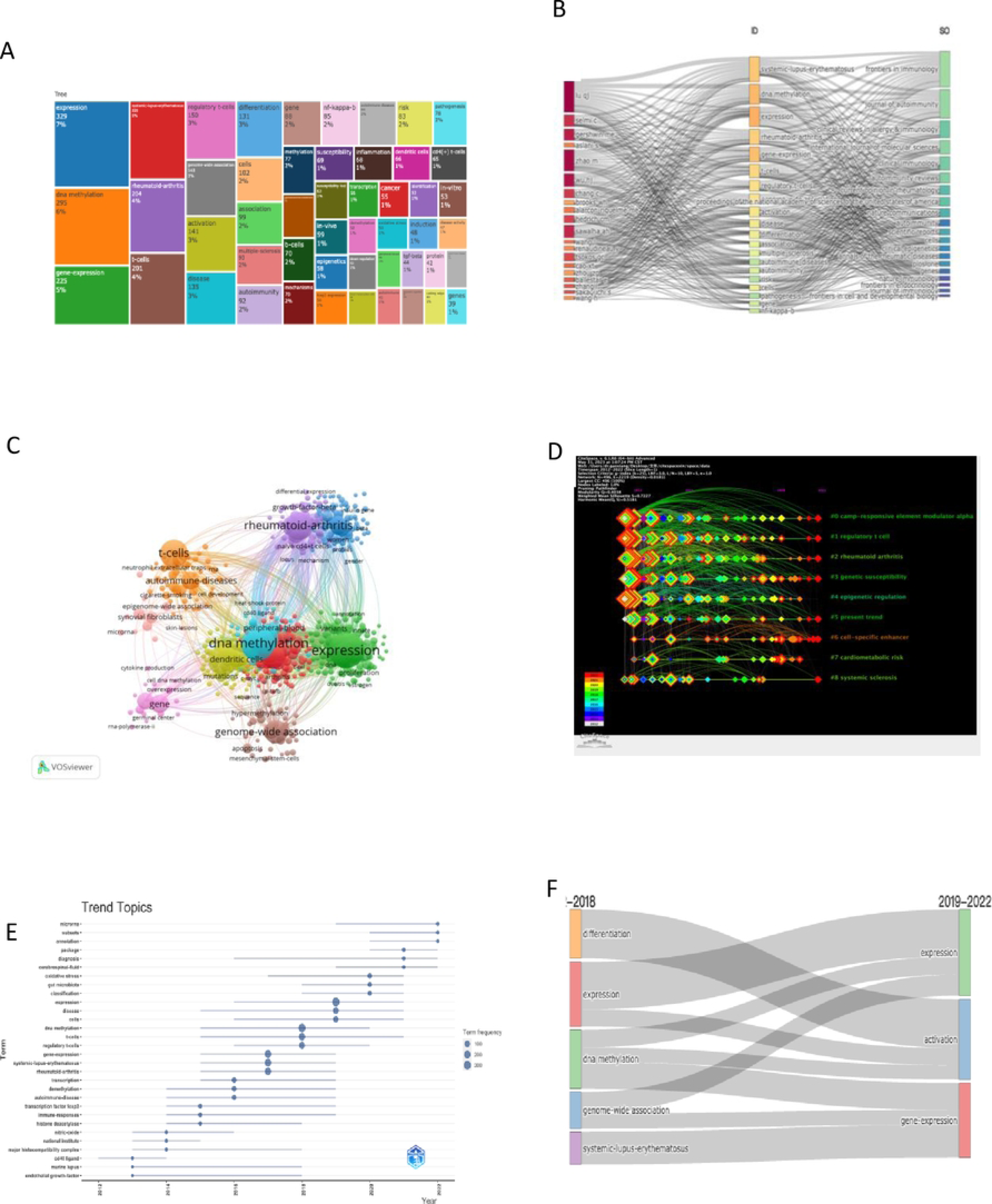

